# Cross-Scanner Reliability of Brain MRI Foundation Models: A Travelling-Heads Study

**DOI:** 10.64898/2026.03.23.26348808

**Authors:** Rafael Navarro-Gonález, Paula Caballero-Lillo, Alessandra Gallo, Santiago Aja-Fernández, Álvaro Planchuelo-Gómez, Rodrigo de Luis-Garía

## Abstract

Foundation models (FMs) are increasingly used for brain MRI analysis, but their reliability across scanners remains unclear. We evaluated seven FMs using 120 acquisitions from 20 participants in the ON-Harmony travelling-heads dataset. We also used a separate cohort of 1,411 healthy adults to train brain-age predictors, either by fitting Ridge regression to frozen embeddings or by fine-tuning the models. We assessed how accurately they predicted age and how consistently their embeddings and age predictions behaved across scanners. AnatCL and y-Aware produced the most reliable embeddings, with distance-based intraclass correlation coefficients (dbICC) of 0.91–0.92, approaching 0.95 for FreeSurfer-derived anatomical measurements. Their shared anatomical preprocessing emerged as a plausible contributor to this pattern. Ridge predictions had higher dbICC than full embeddings for NeuroVFM (0.88 versus 0.19) and Curia-2 (0.84 versus 0.16). After fine-tuning, predictions for the same person differed between scanners by approximately 2 years on average for AnatCL and y-Aware, compared with 4–6 years for NeuroVFM and Curia-2. After correction for multiple comparisons, fine-tuning improved internal-test accuracy in three of seven models and reduced absolute cross-scanner differences in two. These findings favour anatomically focused models for reliable predictions, while highlighting the challenge of preserving useful image information without sacrificing cross-scanner reliability.

## 1 Introduction

Foundation models (FMs) are large neural networks pretrained on broad datasets, often using self-supervised objectives, to learn representations that can be transferred to multiple downstream tasks [1]. Their use has expanded across healthcare, including radiology [2], with applications in brain imaging such as disease classification and anatomical segmentation [3]. An FM encoder converts an image into a numerical feature vector, known as an embedding. For a downstream task, this embedding can be used to train a predictor while keeping the encoder fixed, or the encoder and predictor can be fine-tuned together.

Medical-image representations can retain acquisition signatures, including scanner- and vendor-related information [4]. In structural brain MRI, if scanning the same brain under different acquisition conditions yields substantially different FM-derived outputs, downstream analyses may reflect scanner effects rather than stable participant biology. Crucially, downstream adaptation may not eliminate this nuisance variation: biases encoded in pretrained features can propagate to downstream predictions [5], and scanner sensitivity can remain in predictors trained on frozen FM embeddings [6].

Recent work offers partial evidence. In digital pathology, FM embeddings can carry pronounced scanner-specific signatures [7], and multiple embedding spaces can be organised more strongly by medical centre than by biological factors [8]. Scanner-aware contrastive learning has consequently been proposed to mitigate this sensitivity [6]. In radiology, prior work has assessed same-scanner test–retest embedding similarity in computed tomography (CT) [9, 10], while BrainIAC evaluated downstream prediction robustness in structural brain MRI under synthetic contrast shifts, Gibbs ringing, and bias fields [11]. Whether FM-derived outputs remain stable when the same individuals are scanned across different MRI systems remains unclear.

The travelling-heads paradigm, in which the same participants are scanned under multiple acquisition conditions, provides a direct design for comparing stable participant-associated information with acquisition-related variability [12, 13]. However, measuring the reliability of the full embedding does not tell us whether acquisition differences affect a particular prediction, because a predictor may give little weight to the features that vary across scanners. Fine-tuning changes the representation itself and may therefore alter both prediction accuracy and cross-scanner reliability. Against this background, this study addressed three questions: (1) How reliable are full frozen FM embeddings across acquisition conditions? (2) Are brain-age predictions derived from frozen embeddings reliable across scanners? (3) Does fine-tuning improve predictive accuracy or cross-scanner stability?

We chose brain-age prediction as a widely studied application of machine learning to brain MRI [14, 15]. Models learn age-related imaging patterns from healthy participants and use them to estimate age in new individuals. The difference between predicted and chronological age, often called the brain-age gap, indicates whether a brain appears older or younger than expected relative to those learned patterns. Brain-age differences have been reported across neurological and psychiatric conditions. Brain-age estimates have been associated with progression from mild cognitive impairment to Alzheimer’s disease [16] and with disability progression in multiple sclerosis [17]. Increased brain-age gaps have also been reported in people with chronic migraine [18], psychosis [19], or psychotic experiences [20], among other groups. Its output in years provides an interpretable way to assess both prediction accuracy and consistency across scans of the same person.

In this study, we used the ON-Harmony travelling-heads dataset [21, 22] to benchmark the cross-scanner reliability of frozen embeddings from seven architecturally diverse brain MRI FMs, using FreeSurfer morphometry [23] as a reference. We used a separate cohort of 1,411 healthy adults to develop brain-age predictors using Ridge regression on frozen embeddings or fine-tuning, and evaluated their accuracy in an internal test set. We then applied both approaches to ON-Harmony to assess how consistently they predicted age for the same person across scanners. This design allowed us to examine cross-scanner reliability at each stage, from frozen embeddings to brain-age predictions before and after fine-tuning.

## 2 Methods

### 2.1 Study Design

We evaluated seven FMs: 3D-Neuro-SimCLR [24], AnatCL [25], BrainIAC [11], BrainSegFounder [26], Curia-2 [27], NeuroVFM [28], and y-Aware [29]. Figure 1 summarises the three analyses.

**Fig. 1:**
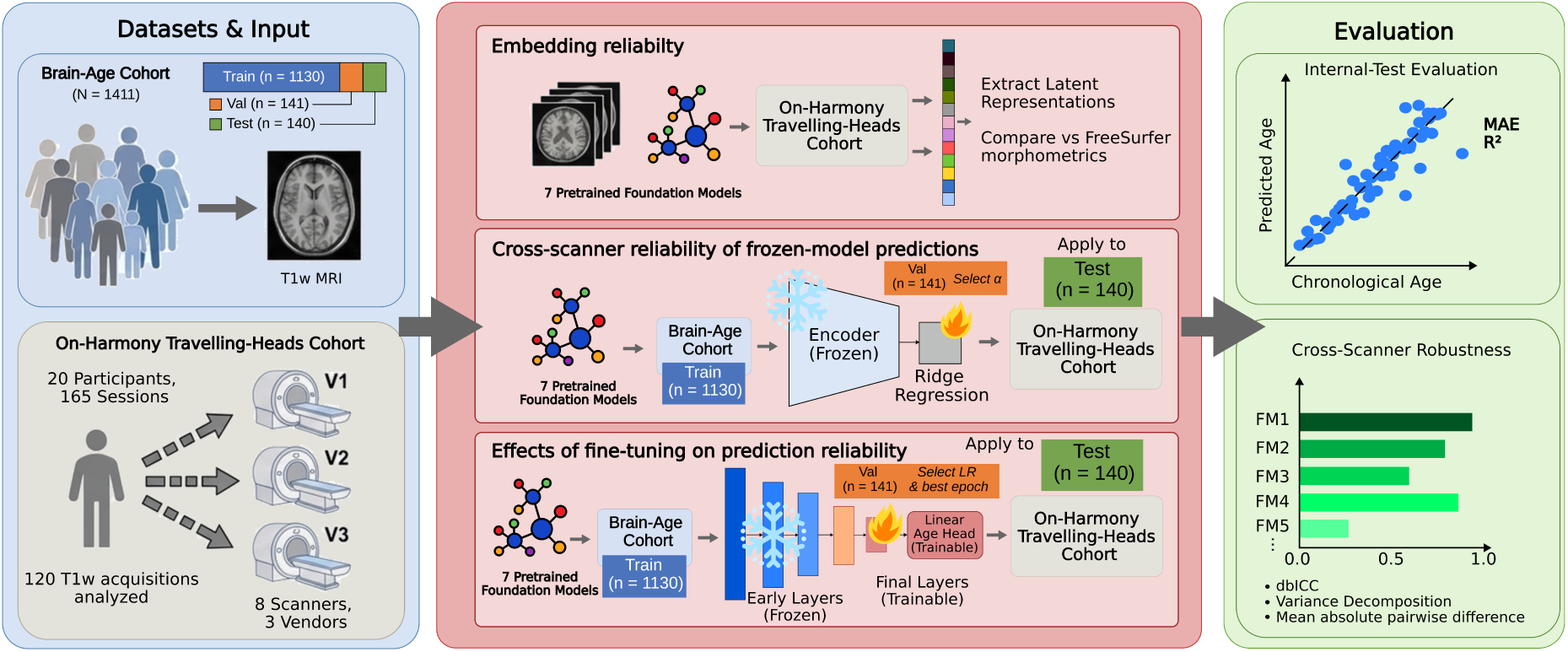
Study workflow for the seven FMs. ON-Harmony provided repeated scans of 20 participants, each scanned on six of the eight scanners. We retained one acquisition per participant–scanner pair (120 scans). A separate cohort of 1,411 healthy adults provided training, validation, and internal-test data for brain-age prediction. First, we measured frozen-embedding reliability, with FreeSurfer morphometry as a reference. Second, we trained Ridge age predictors on frozen embeddings. Third, we fine-tuned each FM’s final encoder block or stage together with an age-prediction head. Both prediction approaches were evaluated for accuracy in the internal test set and for cross-scanner reliability and absolute prediction differences in ON-Harmony, which was excluded from training and model selection.

We began by measuring embedding reliability across scanners in ON-Harmony. We then asked whether brain-age predictions derived from these embeddings were also reliable. We trained Ridge predictors in the separate brain-age development cohort, keeping the encoders frozen. To test whether adapting the FMs changed prediction accuracy or reliability, we fine-tuned the last encoder block or stage together with an age-prediction head. Both prediction approaches were evaluated for accuracy in the internal test set and cross-scanner reliability in ON-Harmony. ON-Harmony was used only for external evaluation, never for training or model selection.

### 2.2 Datasets

#### 2.2.1 ON-Harmony travelling-heads cohort

We used the Oxford-Nottingham Harmonisation (ON-Harmony) dataset (OpenNeuro accession ds004712, version 2.0.1 [21]), a multi-site, multi-modal travelling-heads resource collected between 2018 and 2024. Although the dataset includes multiple MRI modalities, this study analysed only T1-weighted (T1w) structural acquisitions from 20 healthy participants (13 male and 7 female; age 19–50 years, mean *±* standard deviation (SD) 31.50 *±* 10.54 years). The cohort was acquired in two phases. Within each phase, every participant was scanned on six 3T MRI scanners across five imaging sites in Oxford and Nottingham, selected from a total pool of eight systems (Table 1). The T1w acquisition protocols were aligned to the UK Biobank imaging protocol [22]. Four scanners were shared between phases, and two were specific to each phase.

**Table 1:**
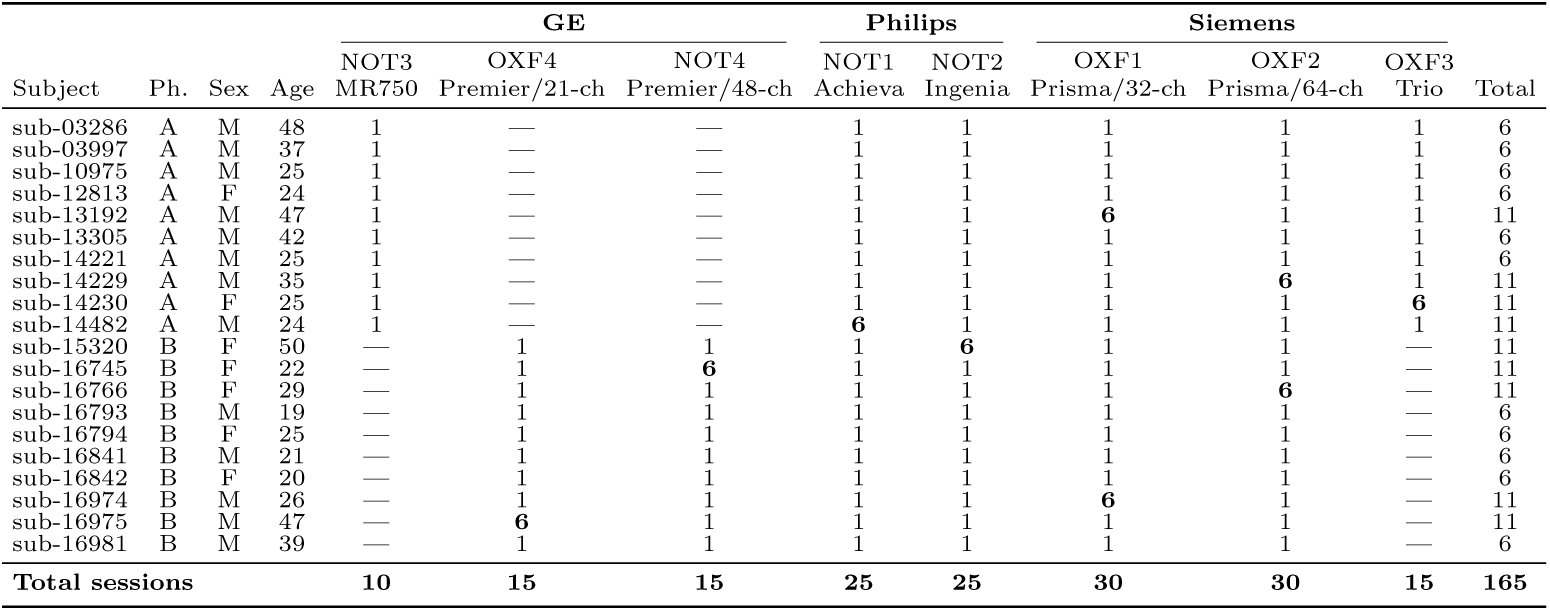
ON-Harmony T1w acquisitions by participant and scanner. Ph. denotes acquisition phase; bold 6 indicates six acquisitions in that cell; — indicates no acquisition; OXF and NOT denote Oxford and Nottingham.

Nine participants had five extra scans on one scanner, bringing the total to 165 acquisitions. For the primary analyses, we used only the first scan from each participant on each scanner (120 acquisitions).

#### 2.2.2 Brain-age development cohort

To assess brain-age prediction using frozen embeddings and fine-tuning, we assembled a separate development cohort from five research datasets. We initially identified 1,427 adults from CamCAN [30], the Neurocognitive aging data release (hereafter NeuroCog) [31], WAND [32], NIMH-RV [33], and MR-ART [34]. CamCAN data were obtained from the Cambridge Centre for Ageing and Neuroscience repository. After exclusions, the final cohort comprised one T1w scan from each of 1,411 healthy adults (794 female and 617 male; age 18–88 years, mean 43.79 *±* 20.55; Table 2).

**Table 2:**
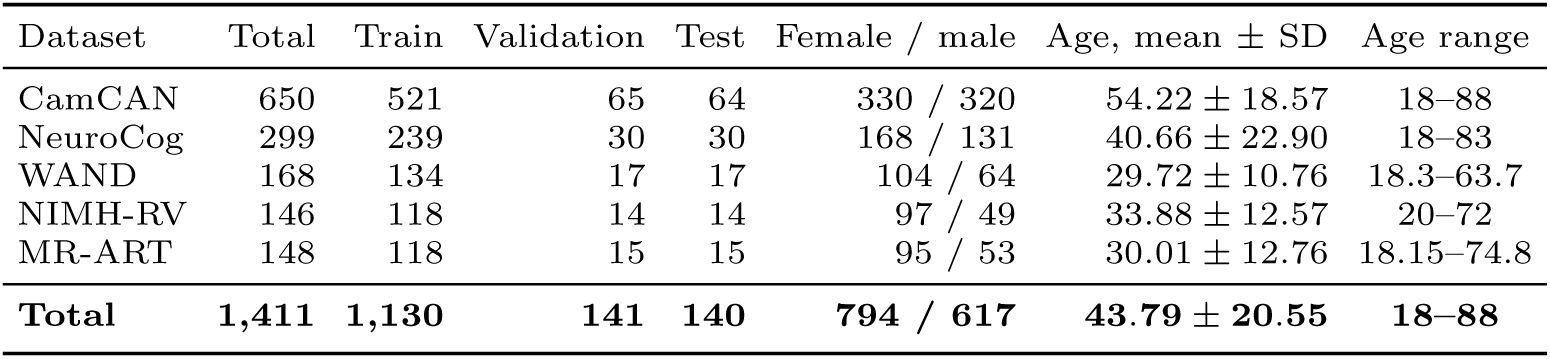
Composition and subject-level partition of the brain-age development cohort. Age and sex summaries include all retained participants within each dataset.

Sixteen participants were excluded before analysis. Nine NIMH-RV participants were marked ineligible, two NeuroCog participants lacked a usable numeric age or sex, and two CamCAN and two WAND candidates had incomplete demographics. One additional CamCAN scan was excluded after two independent failures of the common CAT12/Dartel preprocessing branch. We checked the five datasets against the reported FM pretraining sources and found no shared datasets. Curia-2 and NeuroVFM used private pretraining data, so we could not check for shared participants.

Before model fitting, we generated a deterministic subject-level 80/10/10 split of the 1,411 participants. Allocation was performed within each dataset while preserving the joint distribution of age and sex, yielding 1,130 participants for training, 141 for validation, and 140 for internal testing. Training fitted model parameters, whereas validation alone selected Ridge regularisation, learning-rate configuration, epoch, and neural-network checkpoint. The selected models were evaluated without refitting in the internal test set and in ON-Harmony. Neither was used for training or model selection.

### 2.3 Foundation Models

The seven FMs use convolutional neural networks or Transformers. Five used self-supervised learning (SSL) without participant labels. By contrast, age guided pretraining in both AnatCL and y-Aware, and AnatCL also used anatomical measurements. Below, we describe what each model takes as input and how we obtained one embedding per scan.

1. **3D-Neuro-SimCLR [24]:** This model is a 3D ResNet-18 pretrained on heterogeneous T1w MRI using SimCLR, a contrastive method that brings representations of differently augmented versions of the same image closer together, while separating those of different images [35]. We used a T1w preprocessing pipeline based on TurboPrep [36] and extracted the 512-dimensional global-average-pooled encoder output before the contrastive projection head.
2. **AnatCL [25]:** This model is a 3D ResNet-18 trained on OpenBHB, a brain MRI dataset assembled from multiple cohorts of healthy participants [37]. It used CAT12 voxel-based morphometry (VBM) grey-matter (GM) maps [38] with anatomy- and age-guided contrastive objectives. We used its released fold-0 checkpoint and extracted the 512-dimensional global-average-pooled output before the contrastive head.
3. **BrainIAC [11]:** This model is a 3D ViT-B/16 pretrained with SimCLR on multiparametric brain MRI. We followed the released preprocessing pathway, including N4 bias correction [39], registration, and brain extraction with HD-BET [40], and used the first token of the final 216-token sequence as a 768-dimensional representation. This token represents an image patch rather than a dedicated class (CLS) token.
4. **BrainSegFounder [26]:** This model uses the encoder of SwinUNETR, a segmentation architecture that combines a hierarchical Transformer encoder with a U-shaped decoder [41]. The encoder processes local image regions and progressively combines them into coarser representations covering larger regions. We applied a UK Biobank-derived registered T1w view and spatially averaged its deepest 768-channel feature map.
5. **Curia-2 [27]:** This model is a 2D ViT-B/16 pretrained on CT and MRI using a modified DINOv2 framework [42]. The encoder represents each image patch as a feature vector and adds a learned class (CLS) token to represent the slice as a whole. Through attention, this token can integrate information from the patch tokens. Pretraining uses two complementary objectives: DINO encourages agreement between CLS-based outputs from different views of the same image [43], while iBOT teaches the model to predict representations of masked patches from the visible content [44]. We processed every axial T1w slice with the released processor, extracted its final 768-dimensional CLS representation, and averaged these representations across slices to obtain a volume-level embedding.
6. **NeuroVFM [28]:** This model is a volumetric ViT-B pretrained on clinical CT and MRI using Volumetric Joint-Embedding Predictive Architecture (Vol-JEPA), which adapts the JEPA framework to 3D images [45]. During pretraining, a student encoder processes the visible parts of a volume, and a predictor learns to predict features of the masked patches. A teacher encoder processes the complete volume to provide the target features. The model therefore learns by predicting representations of hidden regions rather than reconstructing their image intensities, without requiring manual labels. Its released preprocessor divides each volume into small 3D patches and removes background patches. The encoder produces a feature vector, or token, for each retained patch, and we averaged the final tokens to obtain a 768-dimensional session representation.
7. **y-Aware [29]:** This model is a 3D DenseNet-121 pretrained on BHB-10K using age-guided contrastive learning and CAT12/VBM GM maps. BHB-10K aggregates 13 healthy MRI cohorts, including the source cohorts used in OpenBHB [37]. We extracted the 1,024-dimensional global-average-pooled encoder output before the projection head.

Their pretraining characteristics are summarised in Table 3, and representative inputs are shown in Supplementary Figure S1. Further details on each model’s preprocessing and representation extraction are provided in the Supplementary Methods and Table S1.

**Table 3:** Pretraining characteristics of the seven foundation models.

| Model | Architecture | Pretraining data | Modalities | Pretraining strategy | Embedding dimension |
|---|---|---|---|---|---|
| 3D-Neuro-SimCLR [24] | 3D ResNet-18 | 44,958 scans; 18,759 patients | T1w | SimCLR contrastive SSL | 512 |
| AnatCL [25] | 3D ResNet-18 | 3,984 scans; 3,984 participants | T1w-derived GM maps | Anatomy- and age-guided contrastive learning | 512 |
| BrainIAC [11] | 3D ViT-B/16 | 32,015 MRI scans | T1w, T2w, FLAIR, T1CE | SimCLR contrastive SSL | 768 |
| BrainSegFounder [26] | 3D SwinUNETR | 82,800 volumes; 41,400 participants | T1w and T2-FLAIR | Reconstruction, rotation and contrastive SSL | 768 |
| Curia-2 [27] | 2D ViT-B/16 | 228 million images | CT and MRI | DINO + iBOT radiology-adapted SSL | 768 |
| NeuroVFM [28] | 3D ViT-B | 5.24 million volumes; 566,915 studies | CT and MRI | Vol-JEPA latent-prediction SSL | 768 |
| y-Aware [29] | 3D DenseNet-121 | 10,420 scans; 7,764 participants | T1w-derived GM maps | Age-guided y-Aware InfoNCE | 1,024 |

AnatCL and y-Aware shared two characteristics: biology-guided pretraining objectives and CAT12/VBM preprocessing. Comparing them with the other five FMs therefore could not establish whether their reliability reflected the training objectives or the anatomically processed inputs. To probe this confounding, we performed a targeted post hoc comparison between AnatCL and AnatCL–SimCLR, the SimCLR baseline released alongside it [25]. The two releases used the same OpenBHB cohort, CAT12/VBM GM maps, 3D ResNet-18 architecture, five-fold design, and 512-dimensional pre-projection readout. They differed in both objective and augmentation policy: AnatCL used age- and anatomy-guided contrastive learning, whereas AnatCL– SimCLR used SimCLR with random cropping and cutout. We compared fold 0 of both releases and repeated the comparison across all five matched folds as a sensitivity analysis.

### 2.4 Analyses

For both cohorts, we applied model-specific preprocessing to the original T1w images supplied by each dataset.

#### 2.4.1 Embedding reliability

We first checked whether the same person’s embeddings remained similar when the scanner changed. We compared frozen FM embeddings with the FreeSurfer (v7.1.0) morphometric features supplied with ON-Harmony [21, 22], using the 120-acquisition sample described above. The same sample was used in all subsequent ON-Harmony analyses.

We measured the reliability of each complete feature vector using the distance-based intraclass correlation coefficient (dbICC) [46]. The dbICC compares differences between scans of the same participant with differences between participants. A high dbICC means that cross-scanner differences within the same participant are small relative to differences between participants. We calculated dbICC on non-standardised feature vectors. As a sensitivity analysis, we examined whether standardising each feature changed the dbICC estimates or model rankings.

To understand where the variability came from, we fitted a random-effects model with participant and scanner random intercepts to each feature. This decomposed its total variance into participant, scanner, and residual components:

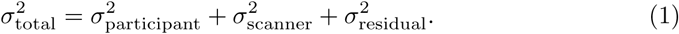

We estimated these components using restricted maximum likelihood (REML). The residual captured variation not explained by participant or scanner effects. We temporarily standardised features to make fitting more stable, then converted the estimated components back to their original scale. Constant coordinates and non-convergent fits were excluded, whereas converged fits with variance components estimated at or near zero were retained. Finally, we summed each component across features and expressed it as a proportion of total variance.

For dbICC, we estimated uncertainty using 1,000 participant-level bootstrap samples. Each sampled participant retained all six selected acquisitions, and paired comparisons used the same participant resamples. To show visually whether embeddings grouped by participant or scanner manufacturer, we also generated two-dimensional projections using t-distributed stochastic neighbour embedding (t-SNE), after standardising each feature.

#### 2.4.2 Cross-scanner reliability of frozen-model predictions

We next checked whether the embeddings could support brain-age predictions that remained consistent across scanners. We used the brain-age development cohort to train Ridge regression models while keeping the FMs unchanged. Ridge predicts age from a weighted combination of embedding features, with a penalty that limits the size of the regression coefficients.

We extracted embeddings with each encoder fixed in evaluation mode. Before fitting Ridge, we standardised each embedding feature and the target age by subtracting their means and dividing by their standard deviations. These statistics were estimated from the 1,130 training participants only and were reused for validation, internal test, and ON-Harmony.

We fitted seven Ridge models with different strengths of regularisation, *α ∈* {0.001, 0.01, 0.1, 1, 10, 100, 1000*}*. Larger values impose a stronger penalty on the coefficients. We converted predictions back to years and selected the value with the lowest mean absolute error (MAE) in the 141-participant validation set. We kept this model as fitted on training data, without refitting on train plus validation, and applied it unchanged to internal test and ON-Harmony. Each FM had one frozen checkpoint and one corresponding Ridge model in the primary analysis. AnatCL used fold 0.

We evaluated three aspects of prediction performance: how closely predicted age matched chronological age, how reliable predictions were across scanners, and how much they differed between scanners in years. We assessed accuracy in the internal test set using MAE and the coefficient of determination (*R*^2^). In ON-Harmony, we assessed cross-scanner reliability using predicted-age dbICC and absolute differences using the mean absolute pairwise difference. As with embeddings, higher dbICC meant that between-participant differences outweighed cross-scanner differences within participants.

To measure the size of cross-scanner differences in years, we calculated the absolute differences between each participant’s six age predictions across all 15 scanner pairs (6 *×* 5*/*2 = 15), then averaged across pairs and participants. This gave the mean absolute pairwise difference in years.

We used 1,000 participant-level bootstrap samples to calculate 95% confidence intervals (CIs). We also checked whether predicting age from frozen embeddings changed cross-scanner reliability. For each FM, we compared the dbICC of its embeddings with that of its Ridge predictions using the same bootstrap samples. This gave seven comparisons, one per FM. We adjusted their two-sided bootstrap *p*-values using the Benjamini–Hochberg (BH) procedure to account for multiple testing.

#### 2.4.3 Effects of fine-tuning on prediction reliability

We then checked whether fine-tuning improved prediction accuracy or consistency across scanners. Using the same brain-age development cohort, we fine-tuned part of each FM together with an age predictor, rather than keeping the encoder fixed as in Ridge. We compared both approaches using the same three measures: internal-test MAE, predicted-age dbICC, and mean absolute pairwise difference across scanners.

We started again from each official checkpoint and kept the encoder frozen except for a prespecified final block or stage. We added a newly initialised linear age-prediction head. This applied the same general strategy across models, but the final architectural units contained different fractions of their parameters (Supplementary Table S2).

We trained the head and the selected encoder parameters using AdamW with a weight decay of 0.01. The loss was mean squared error (MSE), with predicted and true ages expressed on the standardised scale defined by the training set. Participants were processed individually, and their gradients were averaged in groups of eight before each optimiser step. Training ran for at most 30 epochs, with cosine learning-rate annealing and no data augmentation.

For each FM, we first trained three candidate models using seed 0, each with a different learning-rate pair for the encoder and age head: 10*^−^*^6^*/*10*^−^*^4^, 3*×*10*^−^*^6^*/*3*×*10*^−^*^4^, and 10*^−^*^5^*/*10*^−^*^3^. During each run, we measured validation MAE in years after every epoch and saved the model with the lowest value. Training stopped early if validation MAE did not improve by more than 0.01 years for seven consecutive epochs. We then compared the three candidates at their best epochs and selected the learning-rate pair with the lowest validation MAE.

To account for variability between training runs, we repeated training with seeds 1 and 2 while keeping the selected learning rates fixed. For every FM, we calculated each metric separately for the three seeds and averaged the resulting values, following the repeated-run evaluation used by Kumar et al. [47]. For example, we calculated internal-test MAE separately for the three NeuroVFM models and then averaged those MAEs. This mean summarised performance across runs, while the individual results showed how much performance varied. We did not average predicted ages, because our aim was to evaluate fine-tuning rather than construct an ensemble.

In a separate sensitivity analysis for AnatCL, we repeated the frozen-Ridge and fine-tuning analyses across all five pretrained folds to examine whether the checkpoint choice affected the results. Fine-tuning used seed 0. The learning rates selected with fold 0 and seed 0 were reused for the other folds. These fold-level results were kept separate from the primary three-seed averages.

Once selection was complete, we evaluated each retained model on the 140-participant internal test set and on ON-Harmony. We compared each fine-tuned model with its corresponding frozen-Ridge predictor using the same participants and acquisitions, and the same pretrained fold for AnatCL. We expressed these comparisons as gains, with positive values always favouring fine-tuning. For MAE and mean absolute pairwise difference, gain was the frozen value minus the fine-tuned value. For dbICC, it was the fine-tuned value minus the frozen value. Gains were also calculated separately for each seed and then averaged.

To estimate uncertainty in each gain, we used the same participant bootstrap samples for the frozen and fine-tuned models. We calculated paired gain CIs and two-sided bootstrap tests, then applied BH adjustment across the 21 comparisons: seven models for each of the three endpoints, MAE, predicted-age dbICC, and mean absolute pairwise difference. Across all three analyses, we used Python, with PyTorch for embedding extraction and fine-tuning [48] and scikit-learn for Ridge fitting [49]. Random-effects models were fitted in R using lme4 [50].

## 3 Results

### 3.1 Embedding reliability

We first examined whether each person’s frozen representation remained similar across scanners (Figure 2a). Among the seven FMs, AnatCL and y-Aware had the highest dbICC values: 0.92 [0.89, 0.95] and 0.91 [0.84, 0.94], respectively. These approached the FreeSurfer reference of 0.95 [0.92, 0.96]. The additional AnatCL–SimCLR control also had high reliability, with a dbICC of 0.92 [0.89, 0.95]. Its paired comparison with AnatCL and the five-fold sensitivity analysis are reported in Supplementary Table S3. The other FMs had lower reliability. BrainSegFounder had a dbICC of 0.55 [0.53, 0.57], followed by 3D-Neuro-SimCLR, NeuroVFM, Curia-2, and BrainIAC, with estimates ranging from 0.11 to 0.29. Supplementary Table S4 shows how coordinate-wise standardisation changed the dbICC estimates and model rankings.

**Fig. 2:**
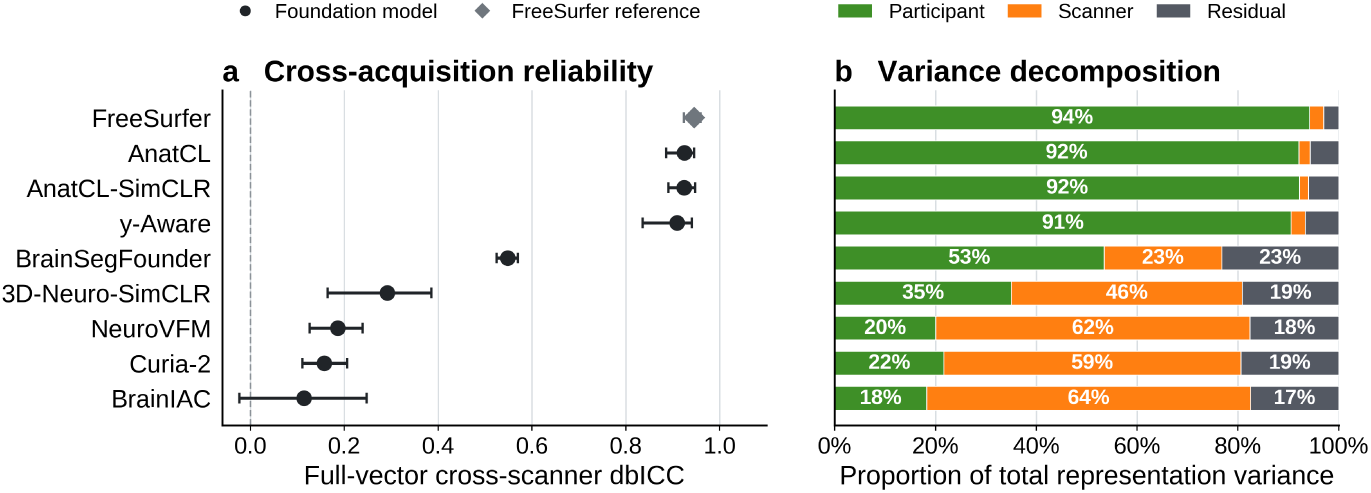
Reliability of the frozen representations using the first acquisition from each participant–scanner cell. **a** Full-vector cross-scanner dbICC; error bars show 95% participant-bootstrap confidence intervals. **b** Participant-, scanner-, and residual-associated proportions obtained by pooling native-scale REML variance-component traces across representation coordinates. Each representation uses one checkpoint, with fold 0 for AnatCL and AnatCL–SimCLR.

We then examined how much of the variation was associated with participants or scanners (Figure 2b). For FreeSurfer, AnatCL–SimCLR, y-Aware, and AnatCL, approximately 90–94% was participant-associated and only 2–3% was scanner-associated. The pattern was different for BrainIAC, NeuroVFM, Curia-2, and 3D-Neuro-SimCLR: scanner-associated variation was the largest component, accounting for approximately 46–64% of the total. BrainSegFounder again lay between these groups, with 53% participant-associated, 23% scanner-associated, and 23% residual variation. Figure 3 shows the corresponding t-SNE projections, coloured by participant and scanner manufacturer.

**Fig. 3:**
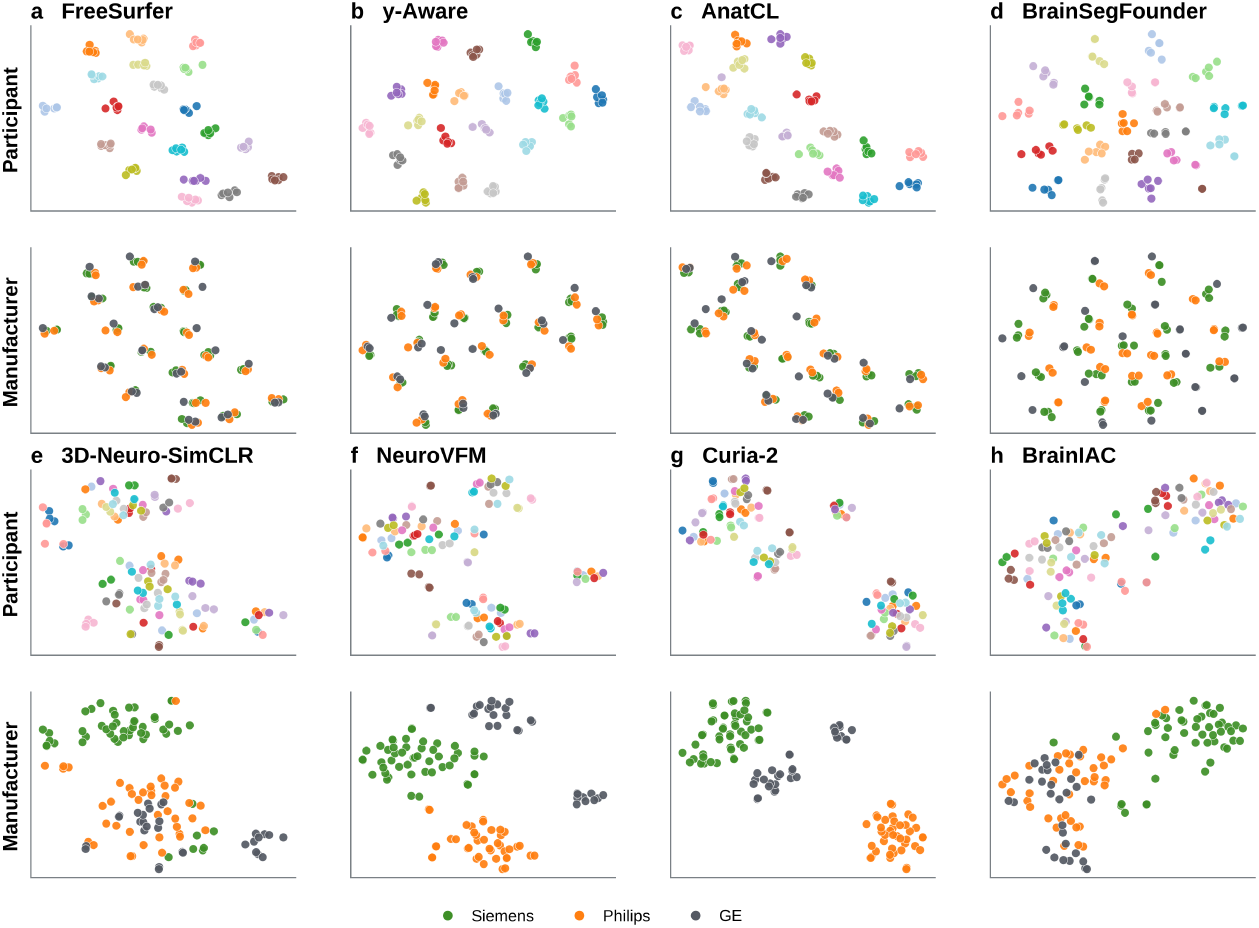
Two-dimensional t-SNE projections of the frozen representations from ON-Harmony for FreeSurfer and the seven foundation models. Each model used the first acquisition from every participant–scanner cell (120 acquisitions). Features were standardised coordinate-wise before t-SNE. Within each model, the upper panel is coloured by participant and the lower panel by scanner manufacturer. AnatCL uses fold 0 because coordinates are not aligned across independently pretrained folds. Axis orientation and scale are arbitrary across models. These projections are descriptive.

### 3.2 Cross-scanner reliability of frozen-model predictions

We next compared embedding reliability with the reliability of Ridge age predictions. For brain-age prediction from frozen embeddings, NeuroVFM had the lowest internal-test MAE at 4.06 years (Figure 4a; Table 4). Curia-2 followed at 4.49 years, while 3D-Neuro-SimCLR and y-Aware both reached 4.54 years. Errors were larger for AnatCL and BrainSegFounder, at 6.34 and 6.83 years, respectively. BrainIAC had the highest MAE, at 11.13 years.

**Fig. 4:**
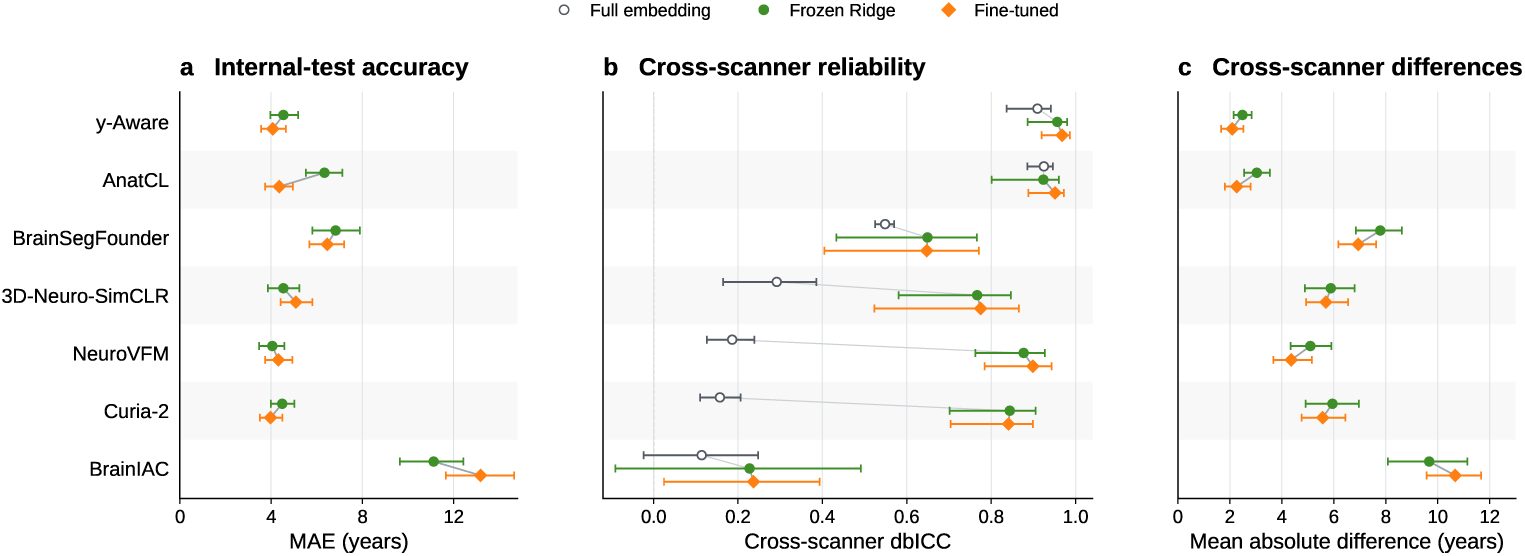
Brain-age prediction with frozen embeddings (green circles, Ridge regression) and after fine-tuning (orange diamonds). **a** Mean absolute error in the 140-participant internal test set. **b** Cross-scanner reliability (dbICC) in ON-Harmony. Open circles show the reliability of the full frozen embedding for comparison. **c** Mean absolute pairwise difference in predicted age between scanners for the same person. Panels b and c use six scans per participant (120 scans in total). Error bars show 95% participant-bootstrap confidence intervals for each estimate.

**Table 4:** Frozen-Ridge and fine-tuned results as estimates [95% participant-bootstrap CIs]. Positive gains favour fine-tuning. Gain CIs use paired bootstraps. Two-sided *p*-values are unadjusted. *q*BH values are adjusted across all 21 comparisons. *^∗^q*BH *<* 0.05. Fine-tuned metrics average three seeds for every model; frozen metrics use one run. AnatCL uses fold 0. ON-Harmony uses the first acquisition per participant–scanner cell.

| Model | Frozen | Fine-tuned | Gain | *p* | *q*<sub>BH</sub> |
|---|---|---|---|---|---|
| **Internal-test MAE (years)** |  |  |  |  |  |
| y-Aware | 4.54 [3.97, 5.19] | 4.08 [3.56, 4.65] | +0.47 [0.14, 0.80] | 0.002 | 0.010<sup>*</sup> |
| AnatCL | 6.34 [5.53, 7.12] | 4.36 [3.74, 4.96] | +1.98 [1.31, 2.66] | 0.002 | 0.010<sup>*</sup> |
| BrainSegFounder | 6.83 [5.81, 7.90] | 6.46 [5.68, 7.21] | +0.37 [−0.37, 1.18] | 0.394 | 0.517 |
| 3D-Neuro-SimCLR | 4.54 [3.87, 5.25] | 5.09 [4.42, 5.81] | −0.55 [−1.04, −0.06] | 0.024 | 0.056 |
| NeuroVFM | 4.06 [3.48, 4.58] | 4.32 [3.74, 4.94] | −0.27 [−0.56, 0.06] | 0.130 | 0.227 |
| Curia-2 | 4.49 [3.99, 5.03] | 3.98 [3.51, 4.49] | +0.51 [0.19, 0.83] | 0.002 | 0.010<sup>*</sup> |
| BrainIAC | 11.13 [9.64, 12.44] | 13.19 [11.67, 14.65] | −2.06 [−3.28, −0.91] | 0.002 | 0.010<sup>*</sup> |
| **ON-Harmony predicted-age dbICC** |  |  |  |  |  |
| y-Aware | 0.96 [0.89, 0.98] | 0.97 [0.92, 0.99] | +0.01 [0.00, 0.04] | 0.016 | 0.048<sup>*</sup> |
| AnatCL | 0.92 [0.80, 0.96] | 0.95 [0.89, 0.97] | +0.03 [0.00, 0.10] | 0.034 | 0.065 |
| BrainSegFounder | 0.65 [0.43, 0.77] | 0.65 [0.40, 0.77] | 0.00 [−0.12, 0.10] | 0.999 | 0.999 |
| 3D-Neuro-SimCLR | 0.77 [0.58, 0.85] | 0.77 [0.52, 0.87] | +0.01 [−0.09, 0.06] | 0.895 | 0.969 |
| NeuroVFM | 0.88 [0.76, 0.93] | 0.90 [0.78, 0.94] | +0.02 [−0.01, 0.06] | 0.194 | 0.291 |
| Curia-2 | 0.84 [0.70, 0.91] | 0.84 [0.70, 0.90] | 0.00 [−0.03, 0.03] | 0.749 | 0.890 |
| BrainIAC | 0.23 [−0.09, 0.49] | 0.24 [0.02, 0.39] | +0.01 [−0.19, 0.27] | 0.923 | 0.969 |
| **ON-Harmony mean absolute pairwise difference (years)** |  |  |  |  |  |
| y-Aware | 2.48 [2.14, 2.84] | 2.09 [1.66, 2.52] | +0.39 [0.12, 0.66] | 0.006 | 0.021<sup>*</sup> |
| AnatCL | 3.04 [2.55, 3.53] | 2.26 [1.81, 2.80] | +0.77 [0.26, 1.35] | 0.004 | 0.017<sup>*</sup> |
| BrainSegFounder | 7.79 [6.85, 8.62] | 6.94 [6.18, 7.63] | +0.85 [0.17, 1.51] | 0.022 | 0.056 |
| 3D-Neuro-SimCLR | 5.88 [4.88, 6.80] | 5.70 [4.94, 6.55] | +0.19 [−0.75, 1.17] | 0.763 | 0.890 |
| NeuroVFM | 5.10 [4.34, 5.91] | 4.36 [3.67, 5.16] | +0.74 [0.11, 1.31] | 0.032 | 0.065 |
| Curia-2 | 5.95 [4.92, 6.97] | 5.57 [4.76, 6.45] | +0.38 [−0.19, 0.94] | 0.216 | 0.302 |
| BrainIAC | 9.67 [8.08, 11.15] | 10.67 [9.57, 11.67] | −1.00 [−2.33, 0.27] | 0.150 | 0.242 |

We next examined whether these age predictions remained consistent across scanners in ON-Harmony. For several models, predicted age was more reliable than the complete embedding (Figure 4b). The largest increases in dbICC were seen for NeuroVFM, from 0.19 to 0.88, Curia-2, from 0.16 to 0.84, and 3D-Neuro-SimCLR, from 0.29 to 0.77. y-Aware showed a smaller increase, from 0.91 to 0.96. All four differences remained statistically significant after BH adjustment. The comparisons for AnatCL, BrainSegFounder, and BrainIAC did not meet the adjusted threshold. Supplementary Table S5 reports all seven differences, their CIs, and their *p* and *q* values.

To express cross-scanner variation in years, we also examined how much predicted age changed for the same person between scanners (Figure 4c). Mean absolute pairwise differences were smallest for y-Aware and AnatCL, at 2.48 and 3.04 years. They ranged from 5.10 to 5.95 years for NeuroVFM, 3D-Neuro-SimCLR, and Curia-2, and reached 7.79 years for BrainSegFounder and 9.67 years for BrainIAC. Individual predictions are shown in Supplementary Figures S2 and S3 for the internal test set and ON-Harmony, respectively.

### 3.3 Effects of fine-tuning on prediction reliability

We compared fine-tuning with frozen-Ridge prediction to see whether it changed accuracy or cross-scanner reliability (Table 4). Supplementary Table S6 reports validation MAE for the three learning-rate pairs, together with the selected pair and best epoch for each model.

Fine-tuning reduced internal-test MAE by 0.47 years for y-Aware, 1.98 years for AnatCL, and 0.51 years for Curia-2 (Figure 4a). These three improvements remained statistically significant after BH adjustment. BrainIAC instead became less accurate, with MAE increasing by 2.06 years. MAE also increased by 0.55 years for 3D-Neuro-SimCLR, but this difference did not meet the adjusted threshold. The changes for BrainSegFounder and NeuroVFM did not meet that threshold either. Results for individual training seeds and the separate five-fold AnatCL sensitivity analysis are provided in Supplementary Table S7.

For cross-scanner reliability, y-Aware was the only model with a statistically significant dbICC increase after adjustment (Figure 4b), with a gain of 0.01. AnatCL’s dbICC increased by 0.03, but this difference did not meet the adjusted threshold. Their predictions also changed less between scanners: the mean absolute pairwise difference decreased by 0.39 years for y-Aware and 0.77 years for AnatCL (Figure 4c). Both reductions remained statistically significant after adjustment. BrainSegFounder and NeuroVFM showed reductions of 0.85 and 0.74 years, but neither met the adjusted threshold. Table 4 gives the estimates, paired gain CIs, and *p* and *q* values for all 21 comparisons. Individual fine-tuned predictions are shown in Supplementary Figures S4 and S5.

## 4 Discussion

We evaluated how consistently seven brain MRI FMs represented the same person across scanners, both through their frozen embeddings and through brain-age predictions obtained with Ridge regression or fine-tuning. Brain-age predictors were developed in a separate cohort and evaluated externally in ON-Harmony. AnatCL and y-Aware produced the most reliable frozen embeddings. Both combine biology-guided pretraining with CAT12 GM inputs, and their dbICC values of 0.92 and 0.91 approached the 0.95 obtained from FreeSurfer morphometric features. The other five FMs had substantially lower values, ranging from 0.11 to 0.55. The variance decomposition supported this pattern: participant-associated variation dominated the most reliable representations, whereas scanner-associated variation dominated four of the five less reliable FMs.

The high embedding reliability of AnatCL and y-Aware carried through to their brain-age predictions, both with frozen-Ridge regression and after fine-tuning. However, reliable full embeddings were not necessary for reliable predictions. NeuroVFM and Curia-2 reached predicted-age dbICC values of 0.88 and 0.84, respectively, despite low full-embedding reliability (dbICC 0.19 and 0.16). AnatCL and y-Aware showed smaller absolute cross-scanner differences with both Ridge and fine-tuning: predictions for the same person differed by approximately 2–3 years between scanners, compared with 4–6 years for NeuroVFM and Curia-2. Thus, high predicted-age dbICC could coexist with substantial cross-scanner differences for individual participants. 3D-Neuro-SimCLR and BrainSegFounder occupied an intermediate position, while BrainIAC showed low reliability at both levels and the largest brain-age prediction errors. An independent brain-age study also reported limited performance for BrainIAC [51].

To our knowledge, this is the first travelling-heads evaluation of FM reliability in neuroimaging. The scanner-associated variation we observed in frozen embeddings is consistent with findings from pathology, where FM representations also retain acquisition-related signatures [7, 8]. Moving to downstream prediction does not necessarily remove this sensitivity. Carloni et al. found scanner-dependent predictions from frozen pathology FM embeddings and reduced these differences using scanner-aware contrastive training [6].

### From anatomical inputs to prediction reliability

AnatCL and y-Aware receive GM maps processed with CAT12 rather than the original MRI images (Supplementary Figure S1). By identifying GM and aligning it to a common anatomical space, this preprocessing may reduce acquisition-related differences while preserving anatomical differences between people. Previous studies provide some support for this explanation. Cole et al. found higher cross-scanner reliability estimates for brain-age predictions based on GM maps than on minimally processed images, although their comparison included only 11 participants [52]. Goto et al. also found that DARTEL registration reduced scanner-related differences in several regional brain volumes [53].

These studies did not test the CAT12 preprocessing used by AnatCL and y-Aware or FM embeddings, but the AnatCL–SimCLR comparison provides additional evidence. It used the same pretraining data, CAT12 inputs, architecture, and readout as AnatCL and also achieved high reliability without biological guidance [25]. The paired comparison provided no evidence of a difference, and the five-fold sensitivity analysis supported the same interpretation (Supplementary Table S3). Although the training recipes also differed in augmentation, this result suggests that shared anatomical preprocessing may help explain the observed reliability. However, scanner differences can persist in CAT12-derived volumes [54].

Architecture and pretraining scale did not provide a simple alternative explanation for full-embedding reliability. AnatCL and 3D-Neuro-SimCLR share a ResNet-18 architecture but had substantially different reliability. NeuroVFM and Curia-2 were pretrained on much larger datasets than AnatCL, yet their embeddings were less reliable across scanners. These comparisons do not isolate the effects of architecture or dataset size, but they show that neither a shared architecture nor larger pretraining datasets guarantee similar or higher reliability.

The picture changed when we used the frozen embeddings to predict age with Ridge regression. The increase in reliability for NeuroVFM and Curia-2 (Supplementary Table S5) may reflect how Ridge combines their features. It learns a weighted combination for predicting age, so features that vary across scanners may contribute relatively little to the prediction. A task-specific readout can therefore be more reliable without making the full embedding reliable. However, the same flexibility also allows a predictor to use acquisition-related features as clues to age when acquisition and age are linked in the training data. This form of shortcut learning [55] cannot be ruled out by high prediction reliability.

These gains in reliability also need to be distinguished from agreement across scanners and accuracy against chronological age. A high dbICC means that differences between participants are large relative to differences between their scans, not that those scans yield nearly identical predictions. Nor does it establish accuracy: a predictor could consistently underestimate age across all scanners. MAE describes error against chronological age, whereas the mean absolute pairwise difference describes how much predictions change for the same person between scanners. Together with dbICC, these measures answer distinct questions about whether a brain-age prediction can serve as a useful individual measurement.

### Brain-age utility and fine-tuning

Fine-tuning did not provide a general solution to either prediction error or scanner sensitivity (Table 4). Some models became more accurate, but improvements in cross-scanner reliability were less consistent. This distinction matters because training minimised age-prediction error, not differences between scans of the same person. Adapting the encoder may strengthen useful age-related features, but it can also favour acquisition-related patterns correlated with age or alter useful pretrained features [47]. A model selected for lower validation MAE may therefore remain sensitive to scanner changes. The starting points also differed: age information was already used during y-Aware and AnatCL pretraining, so brain-age prediction was not an equally new task for every model.

After fine-tuning, AnatCL and y-Aware combined high cross-scanner reliability with internal-test MAEs of approximately four to five years, similar to Curia-2 and NeuroVFM. Their GM maps therefore retained enough information for competitive brain-age prediction, despite excluding much of the original image content. More image information did not translate into a consistent advantage under our fine-tuning protocol. However, information outside these maps may still be useful for other tasks or with more extensive adaptation. Wang et al. have described a related trade-off: reducing differences between imaging conditions can also remove useful predictive information [56]. The challenge is to make use of that information without relying on acquisition-related cues.

BrainIAC’s poor performance also prompted us to explore more aggressive fine-tuning. Higher encoder learning rates reduced validation MAE but showed greater overfitting, with a wider gap between training and validation loss (Supplementary Figure S6).

Averaging metrics across training seeds reduced dependence on any single run. However, similar internal-test MAEs across seeds did not always imply similar cross-scanner reliability, particularly for BrainSegFounder and 3D-Neuro-SimCLR (Supplementary Table S7). Table 5 summarises the main findings for each model across embedding reliability, brain-age prediction, and fine-tuning.

**Table 5:** Summary of findings across the seven foundation models. Descriptions compare the evaluated models, not clinical categories. Prediction error refers to internal-test MAE, with lower values indicating better performance. Prediction consistency refers to ON-Harmony dbICC and mean absolute pairwise differences across scanners. Fine-tuning conclusions account for the global multiple-comparison adjustment. Findings apply to the inputs, readouts and adaptation protocol used here. Numerical estimates are reported in Table 4 and Figures 2 and 4.

| **Model** | **Embedding reliability** | **Frozen-Ridge prediction error** | **Frozen prediction consistency** | **Effect of fine-tuning** |
|---|---|---|---|---|
| **y-Aware** | Higher. | Lower. | High dbICC and comparatively small cross-scanner differences. | Reduced prediction error, increased dbICC and improved absolute agreement. |
| **AnatCL** | Higher. | Intermediate. | High dbICC and comparatively small cross-scanner differences. | Largest reduction in prediction error and improved absolute agreement. No clear dbICC gain after correction. |
| **BrainSeg Founder** | Intermediate. | Intermediate. | Intermediate dbICC and large cross-scanner differences. | No clear benefit after correction. |
| **3D-Neuro-SimCLR** | Lower. | Lower. | Higher dbICC than its embeddings, but large cross-scanner differences. | Prediction error increased with borderline evidence. No clear cross-scanner benefit. |
| **NeuroVFM** | Lower. | Lower. | High dbICC, but cross-scanner differences of several years. | No clear benefit after correction. |
| **Curia-2** | Lower. | Lower. | High dbICC, but large cross-scanner differences and marked external age bias. | Reduced prediction error. No clear cross-scanner benefit. |
| **BrainIAC** | Lower. | Higher. | Low dbICC and large cross-scanner differences. | Increased prediction error. No clear cross-scanner benefit. |

### Limitations

Our study has several limitations. ON-Harmony included only 20 healthy adults aged 19–50, which limits the precision of our estimates. dbICC also depends on differences between participants, so estimates from this relatively homogeneous sample may be lower than those from a more diverse population [46]. Each participant was scanned on six systems within their acquisition phase, but only four scanners were shared between phases. The pairwise-distance estimator can be computed from these observed acquisitions without requiring every participant–scanner combination. However, the different scanner sets may influence the within- and between-participant distances and therefore the dbICC estimates. To examine this, we repeated the analysis using only the four shared scanners, retaining all 20 participants and 80 acquisitions. The broad separation between models persisted, although individual estimates changed. For example, BrainSegFounder’s dbICC increased from 0.55 to 0.64 (Supplementary Table S8). The scanner-associated variance estimated by REML reflects the acquisition conditions as a whole, not hardware alone.

Each FM combined its own preprocessing, architecture, and pretraining strategy, so we could not isolate their individual contributions. The same fine-tuning rule also updated different proportions of each encoder: approximately 8% in the ViT-based models and 75% in the ResNet and Swin models, with y-Aware between these extremes (Supplementary Table S2). Furthermore, using the released checkpoints did not always provide a complete route from an MRI scan to an embedding. Some implementation details were unclear or differed between the papers and public code, so we made study-specific decisions. In particular, several models required us to choose how to combine their outputs into a single vector for the whole volume (Supplementary Table S1). Different choices at this step could change the measured reliability.

Curia-2 systematically underestimated age in ON-Harmony: its frozen-Ridge mean error shifted from *−*0.18 years in the internal test set to *−*14.08 years externally (Supplementary Figure S3f). The cause of this external calibration shift remains unclear. Differences in acquisition conditions and their interaction with the slice-wise preprocessing and volumetric readout may have contributed, but our analyses did not isolate their effects.

The FreeSurfer reference also requires care in interpretation because its reliability depends on how the feature vector is defined. Its vector combines volumes, surface areas, and thicknesses with different units and scales (Supplementary Table S9). Because our primary dbICC uses squared Euclidean distances without coordinate-wise standardisation, features with greater numerical variation contribute more. This sensitivity also affected learned embeddings: after coordinate-wise standardisation, y-Aware’s dbICC decreased from 0.91 to 0.78, whereas AnatCL’s decreased from 0.92 to 0.89. Feature selection or rescaling can therefore change the estimate and ranking (Supplementary Table S4).

Furthermore, the brain-age cohort reflects a realistic multi-cohort setting, with unequal age and acquisition distributions across five datasets. This heterogeneity also means that age, dataset, and acquisition were partly confounded, potentially allowing scanner-related shortcuts. Identical subject-level splits support fair model comparisons, and ON-Harmony provides a separate external evaluation, but neither removes this confounding. Finally, our cross-scanner findings apply to 3T T1-weighted MRI and the evaluated frozen-Ridge and fine-tuning protocols. Other modalities, tasks, adaptation depths, or objectives may behave differently.

## 5 Conclusion

In this travelling-heads study, AnatCL and y-Aware produced the most reliable embeddings among the seven FMs, approaching the FreeSurfer morphometric reference. The similarly high reliability of AnatCL–SimCLR suggests that shared anatomical preprocessing may contribute to this pattern.

When frozen embeddings were used to predict brain age with Ridge regression, AnatCL and y-Aware also produced predictions with high cross-scanner dbICC. NeuroVFM and Curia-2 produced substantially more reliable age predictions than their full embeddings, showing that a task-specific predictor can recover stable information from a less reliable representation. However, their predictions for the same person still varied more across scanners than those of AnatCL and y-Aware. Fine-tuning did not consistently improve accuracy or cross-scanner stability. After adaptation, AnatCL and y-Aware combined high cross-scanner reliability with brain-age prediction errors comparable to Curia-2 and NeuroVFM.

Overall, anatomically focused models combined competitive brain-age prediction with high cross-scanner reliability. Other models produced age predictions that were much more reliable than their full embeddings, but still varied more between scanners for the same person.

## Declarations

### Ethics approval

This study analysed existing, de-identified data from ON-Harmony, CamCAN, NeuroCog, WAND, NIMH-RV, and MR-ART. Ethical approvals and participant consent for the original data collection and sharing are described in the respective dataset publications. No new participants were recruited for this study.

### Supplementary Material

Supplementary Methods, Tables, and Figures are provided in the accompanying supplementary document.

### Use of generative AI

OpenAI Codex was used to assist with manuscript drafting and editing, code development, debugging, and review. The authors reviewed the resulting text and code and take full responsibility for the analyses, interpretation, and final manuscript.

### Author contributions (CRediT)

**R.N.-G.:** Conceptualization, Methodology, Software, Formal analysis, Investigation, Data curation, Writing (original draft), Visualization. **P.C.-L.:** Investigation, Validation, Writing (review & editing). **A.G.:** Software, Validation, Writing (review & editing). **S.A.-F.:** Writing (review & editing). **A.P.-G. and R.d.L.-G.:** Conceptualization, Writing (review & editing), Funding acquisition.

## Data Availability

This study used existing MRI data from six datasets. ON-Harmony (version 2.0.1) is available at https://openneuro.org/datasets/ds004712/versions/2.0.1. The brain-age development datasets were NeuroCog (https://openneuro.org/datasets/ds003592 accompanying demographics: https://osf.io/yhzxe/), WAND (https://doi.org/10.12751/g-node.5mv3bf), NIMH-RV (https://openneuro.org/datasets/ds004215), MR-ART (https://openneuro.org/datasets/ds004173), and CamCAN. CamCAN data are available upon approved application through the Cambridge Centre for Ageing and Neuroscience repository (https://opendata.mrc-cbu.cam.ac.uk/projects/camcan/) under its data-use agreement.
Analysis code and derived data, including embeddings and brain-age predictions, are available at https://github.com/rafaloz/Reliability_FMs. Source MRI scans and participants chronological ages are not redistributed in this repository and must be obtained from the original data sources.

https://openneuro.org/datasets/ds004712

https://github.com/rafaloz/Reliability_FMs

https://openneuro.org/datasets/ds003592

https://doi.org/10.12751/g-node.5mv3bf

https://openneuro.org/datasets/ds004215

https://openneuro.org/datasets/ds004173

https://opendata.mrc-cbu.cam.ac.uk/projects/camcan/

## Acknowledgements

We thank the participants and teams who collected and shared the six datasets, including the ON-Harmony consortium. CamCAN provided data collection and sharing, funded by the UK Biotechnology and Biological Sciences Research Council (BB/H008217/1), with additional support from the UK Medical Research Council and the University of Cambridge, UK.

## Data availability

ON-Harmony version 2.0.1 is available at https://openneuro. org/datasets/ds004712/versions/2.0.1. The development datasets are available through the CamCAN repository (https://opendata.mrc-cbu.cam.ac.uk/projects/camcan/), NeuroCog (https://openneuro.org/datasets/ds003592; accompanying demographic and behavioural data at https://osf.io/yhzxe/), WAND (https://doi.org/10.12751/g-node.5mv3bf), NIMH-RV (https://openneuro.org/datasets/ds004215), and MR-ART (https://openneuro.org/datasets/ds004173). ON-Harmony, NeuroCog, NIMH-RV, and MR-ART are distributed under CC0. WAND is distributed under CC BY 4.0 (https://creativecommons.org/licenses/by/4.0/). CamCAN access requires an approved application and compliance with its data-use agreement; source data are not redistributed with our code.

## Code availability

Code and analysis scripts are publicly available on GitHub at https://github.com/rafaloz/ReliabilityFMs.

