## Supplementary for "Cross-Scanner Reliability of Brain MRI Foundation Models: A Travelling-Heads Study"

| Model | Model input | Spatial preprocessing | Intensity processing | Encoder output and session aggregation | Dim. |
|---|---|---|---|---|---|
| 3D-Neuro-SimCLR [1] | T1w and derived brain mask | N4 correction [2]; SynthStrip [3]; rigid registration to the complete ICBM152 2009c nonlinear symmetric template [4]; SynthSeg-derived mask [5]; transpose and centre crop to 150 × 192 × 192 | Negative values clamped to zero after skull stripping; WhiteStripe normalisation [6]; masked z-score at model loading | ResNet-18 GAP output before the 512 → 512 → 64 contrastive projector; one vector per volume | 512 |
| AnatCL [7] | CAT12 modulated grey-matter (GM) map | CAT12 tissue segmentation and Dartel normalisation [8] with the IXI555 MNI152 template at 1.5-mm isotropic resolution; 121 × 145 × 121 map cropped to 121 × 128 × 121 | CAT12 GM intensities retained; no additional normalisation | ResNet-18 GAP output before the 512 → 512 → 128 contrastive head; fold 0 primary, other folds evaluated separately | 512 |
| BrainIAC [9] | T1w | N4 correction [2]; rigid registration to the released temp_head atlas at 1-mm resolution; HD-BET [10]; trilinear resize to $96^3$ | Z-score over nonzero voxels | First token after the final normalisation of the 216-token 3D ViT sequence. The model has no learned CLS token; the first-patch readout follows the released inference path | 768 |
| BrainSeg Founder [11] | T1w | fslreorient2std; robustfov 170-slice crop; BET; standard-space ROI generation; FLIRT and FNIRT to MNI152 1-mm space; warp composition and masking [12]; RAS foreground crop, padding, and trilinear resize to $96^3$ | Nonzero-voxel z-score before resizing | Spatial mean of the deepest Swin bottleneck, $[768, 3, 3, 3] \to [768]$; the segmentation decoder and SSL heads were excluded | 768 |
| Curia-2 [13] | Native T1w axial slices | Volume reorientation to LAS; every axial slice rotated from LA to PL and resized bicubically to 512 × 512; no skull stripping or registration | Lower clamp at $-1000$ followed by z-score over the complete slice | Final-layer learned CLS token for each slice, averaged with equal weight across all axial slices; no inter-slice attention | 768 |
| NeuroVFM [14] | Native T1w; foreground patches | Released preprocessor: RPI orientation; acquisition-aware resampling to 1 mm in-plane and 4 mm through-plane; central crop to dimensions divisible by 4 × 16 × 16; foreground patch selection | 0.5th–99.5th percentile clipping, min–max scaling, and released MRI input normalisation | Mean of all variable-length, final-normalised 4 × 16 × 16 foreground-patch tokens; no truncation or learned pooling | 768 |
| y-Aware [15] | CAT12 modulated GM map | CAT12 tissue segmentation and Dartel normalisation [8] with the IXI555 MNI152 template at 1.5-mm isotropic resolution; complete 121 × 145 × 121 map retained | Whole-volume z-score | DenseNet-121 GAP output before the 1024 → 512 → 128 contrastive projector | 1,024 |

**Table S1**: Model-specific preprocessing and extraction used for the primary representations. GAP denotes global average pooling. Operations are listed in execution order; model-specific implementation qualifications are described below.

We followed the released preprocessing pipelines, with a few implementation choices. For 3D-Neuro-SimCLR, we set negative background intensities to zero after SynthStrip and before registration. For BrainIAC, we used the released preprocessing without adding denoising, as the release did not clearly specify this step. For NeuroVFM, we followed the public code, which selects foreground using a 10th-percentile threshold, rather than the Otsu method described in the paper.

For BrainSegFounder, we used the single-channel UK Biobank Stage 1 checkpoint. We followed its UK Biobank-derived preprocessing but omitted gradient-distortion correction because scanner-specific coefficient files were unavailable. Resizing the input to $96^3$ and averaging the deepest feature map were study-specific choices.

The results also depend on how encoder outputs were converted into one vector per scan. We used BrainIAC's first patch token, BrainSegFounder's spatial mean, Curia-2's mean across axial-slice CLS tokens, and NeuroVFM's mean across foreground tokens. Our findings apply to these representations. We visually reviewed the processed inputs for anatomical coverage, orientation, registration failures, and image corruption.

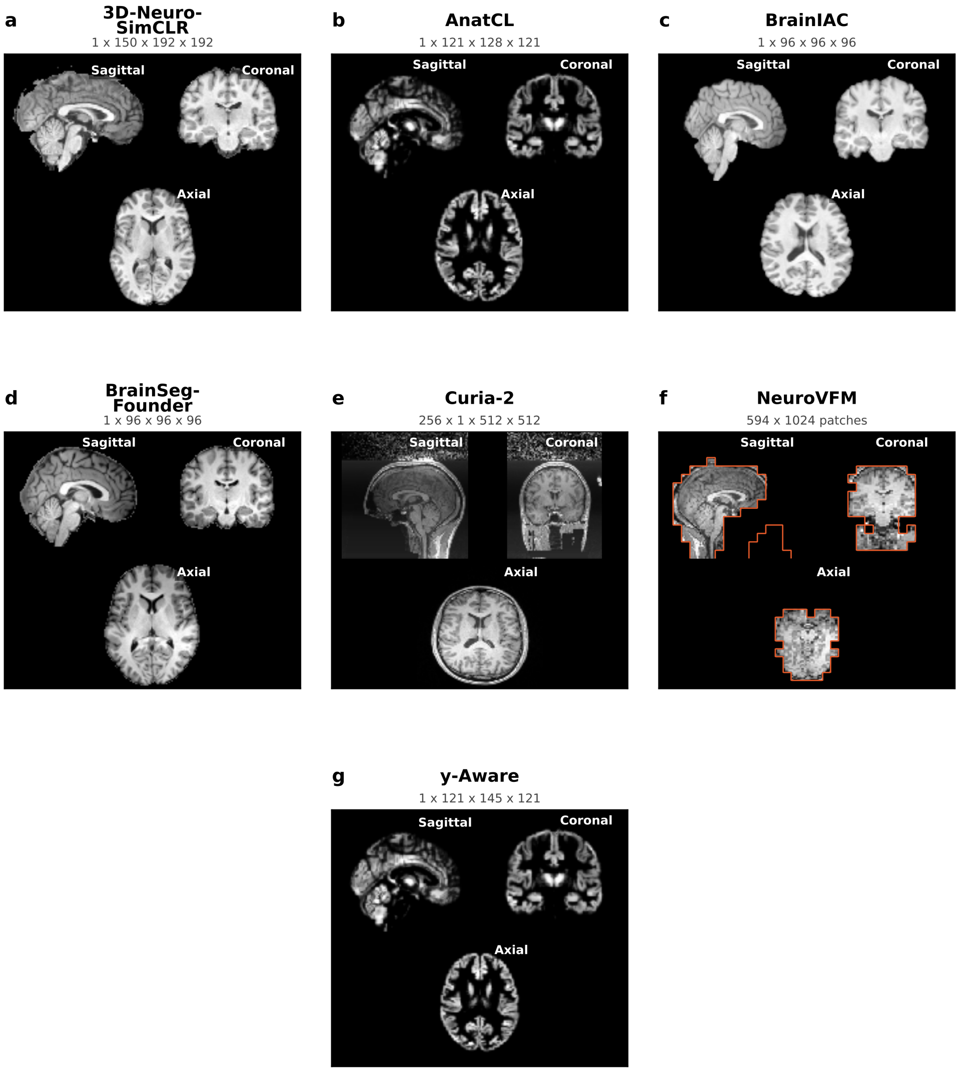


**Fig. S1**: Inputs to the seven FMs for the same participant (CamCAN, sub-CC110033), shown in central sagittal, coronal, and axial views. **a** 3D-Neuro-SimCLR: registered, masked, and standardised T1w image. **b** AnatCL: cropped CAT12 GM map. **c** BrainIAC: registered, skull-stripped, and normalised T1w image. **d** BrainSegFounder: T1w image after UK Biobank-derived preprocessing. **e** Curia-2: each axial slice is processed independently. The sagittal and coronal views were reconstructed by stacking these slices for display. **f** NeuroVFM: views reconstructed from cached foreground patches, before MRI input normalisation. Orange outlines mark the retained patches; discarded background patches appear black. **g** y-Aware: complete CAT12 GM map after whole-volume standardisation.

| Model | Trainable encoder component | Encoder trainable (%) |
|---|---|---|
| 3D-Neuro-SimCLR | Final 3D ResNet stage (layer4) | 75.1 |
| AnatCL | Final 3D ResNet stage (layer4) | 75.1 |
| BrainIAC | Final Transformer block and final LayerNorm | 8.0 |
| BrainSegFounder | Final Swin stage, including its terminal patch merging | 74.7 |
| Curia-2 | Final DINOv2 block and final LayerNorm | 8.2 |
| NeuroVFM | Final volumetric ViT block and final LayerNorm | 8.3 |
| y-Aware | Final DenseNet block (denseblock4) | 29.7 |

**Table S2**: Fine-tuning boundaries. Percentages refer to the active pretrained encoder and exclude the newly initialised linear age head and any unused pretraining heads or decoder. Encoders remained in evaluation mode during fine-tuning, with BatchNorm running statistics fixed where present.

| Analysis | AnatCL dbICC | AnatCL–SimCLR dbICC | Difference [95% CI] | $p$ |
|---|---|---|---|---|
| Fold 0 | 0.92 [0.89, 0.95] | 0.92 [0.89, 0.95] | 0.00 [−0.04, 0.04] | 0.96 |
| Five-fold sensitivity | 0.90 [0.88, 0.92] | 0.92 [0.89, 0.94] | −0.02 [−0.05, 0.01] | 0.18 |

**Table S3**: AnatCL and AnatCL–SimCLR embedding reliability in the primary fold-0 comparison and the five-fold sensitivity analysis. Values are dbICC [95% CI]. Differences are AnatCL minus AnatCL–SimCLR. Five-fold estimates average independently calculated fold-level metrics within each shared participant bootstrap, not embeddings. Both comparisons use the same 120 acquisitions and 1,000 participant-bootstrap samples. Two-sided $p$-values are reported separately from the brain-age multiple-comparison families.

| Representation | Dimensions | Native features dbICC | Native features Rank | Standardised features dbICC | Standardised features Rank |
|---|---|---|---|---|---|
| FreeSurfer | 201 | 0.95 | 1 | 0.87 | 3 |
| AnatCL | 512 | 0.92 | 2 | 0.89 | 2 |
| AnatCL–SimCLR | 512 | 0.92 | 3 | 0.89 | 1 |
| y-Aware | 1024 | 0.91 | 4 | 0.78 | 4 |
| BrainSegFounder | 768 | 0.55 | 5 | 0.55 | 5 |
| 3D-Neuro-SimCLR | 512 | 0.29 | 6 | 0.25 | 6 |
| NeuroVFM | 768 | 0.19 | 7 | 0.24 | 7 |
| Curia-2 | 768 | 0.16 | 8 | 0.20 | 8 |
| BrainIAC | 768 | 0.11 | 9 | 0.14 | 9 |

**Table S4**: Post hoc sensitivity of full-vector dbICC and rankings to coordinate-wise scaling. Native values reproduce the primary analysis. Standardised values use zero-mean, unit-variance coordinates, with scaling estimated separately for each representation and fold from the same 120 selected ON-Harmony acquisitions (20 participants).

| Model | Embedding dbICC | Ridge dbICC | Difference [95% CI] | $p$ | $q_{\text{BH}}$ |
|---|---|---|---|---|---|
| y-Aware | 0.91 | 0.96 | +0.05 [0.02, 0.08] | < 0.01 | 0.01* |
| AnatCL | 0.92 | 0.92 | 0.00 [−0.11, 0.04] | 0.93 | 0.93 |
| BrainSegFounder | 0.55 | 0.65 | +0.10 [−0.11, 0.22] | 0.32 | 0.41 |
| 3D-Neuro-SimCLR | 0.29 | 0.77 | +0.47 [0.34, 0.59] | < 0.01 | < 0.01* |
| NeuroVFM | 0.19 | 0.88 | +0.69 [0.59, 0.75] | < 0.01 | < 0.01* |
| Curia-2 | 0.16 | 0.84 | +0.69 [0.55, 0.77] | < 0.01 | < 0.01* |
| BrainIAC | 0.11 | 0.23 | +0.11 [−0.10, 0.27] | 0.35 | 0.41 |

**Table S5**: Cross-scanner reliability of frozen embeddings and their Ridge age predictions in the same 120 ON-Harmony acquisitions. Differences are prediction minus embedding dbICC. Confidence intervals and two-sided $p$-values use 1,000 paired participant-bootstrap samples. The $q_{\text{BH}}$ values apply BH adjustment across these seven comparisons.

| Model | Validation MAE (years) | | | Selected configuration | |
|---|---|---|---|---|---|
| | Conservative | Intermediate | High | LR encoder/head | Epoch |
| y-Aware | 4.337 | 3.898 | **3.615** | $10^{-5}/10^{-3}$ | 27 |
| AnatCL | 6.601 | 5.507 | **4.452** | $10^{-5}/10^{-3}$ | 26 |
| BrainSegFounder | 13.129 | 9.338 | **5.996** | $10^{-5}/10^{-3}$ | 27 |
| 3D-Neuro-SimCLR | 6.038 | **5.739** | 5.740 | $3 \times 10^{-6}/3 \times 10^{-4}$ | 18 |
| NeuroVFM | 10.258 | 4.612 | **4.275** | $10^{-5}/10^{-3}$ | 23 |
| Curia-2 | 6.179 | 4.325 | **3.373** | $10^{-5}/10^{-3}$ | 25 |
| BrainIAC | 12.762 | 12.418 | **11.882** | $10^{-5}/10^{-3}$ | 26 |

**Table S6**: Validation MAE for the three fine-tuning learning-rate (LR) pairs: conservative ($10^{-6}/10^{-4}$, encoder/head), intermediate ($3\times10^{-6}/3\times10^{-4}$), and high ($10^{-5}/10^{-3}$). Bold identifies the selected configuration. Each MAE uses the candidate's best validation epoch. Selection used unrounded MAEs, seed 0 and, for AnatCL, fold 0. The selected LR pair and epoch are shown on the right.

**A. Fine-tuning by training seed**

| | Internal-test MAE (years) | | | ON-Harmony dbICC | | | Mean absolute pairwise difference (years) | | |
|---|---|---|---|---|---|---|---|---|---|
| Model | Seed 0 | Seed 1 | Seed 2 | Seed 0 | Seed 1 | Seed 2 | Seed 0 | Seed 1 | Seed 2 |
| y-Aware | 4.02 | 4.02 | 4.19 | 0.97 | 0.97 | 0.97 | 2.10 | 2.08 | 2.09 |
| AnatCL (fold 0) | 4.37 | 4.38 | 4.32 | 0.95 | 0.95 | 0.95 | 2.24 | 2.27 | 2.28 |
| BrainSegFounder | 6.42 | 6.44 | 6.52 | 0.68 | 0.68 | 0.58 | 6.66 | 6.25 | 7.92 |
| 3D-Neuro-SimCLR | 5.02 | 5.10 | 5.16 | 0.73 | 0.78 | 0.81 | 6.50 | 5.61 | 4.98 |
| NeuroVFM | 4.30 | 4.32 | 4.36 | 0.90 | 0.90 | 0.90 | 4.35 | 4.40 | 4.32 |
| Curia-2 | 3.99 | 3.94 | 4.00 | 0.85 | 0.84 | 0.83 | 5.22 | 5.72 | 5.77 |
| BrainIAC | 13.18 | 13.17 | 13.21 | 0.23 | 0.24 | 0.24 | 10.50 | 10.68 | 10.83 |

**B. AnatCL by pretraining fold**

| | Internal-test MAE (years) | | ON-Harmony dbICC | | Mean absolute pairwise difference (years) | |
|---|---|---|---|---|---|---|
| Fold | Frozen | Fine-tuned | Frozen | Fine-tuned | Frozen | Fine-tuned |
| 0 | 6.34 | 4.37 | 0.92 | 0.95 | 3.04 | 2.24 |
| 1 | 6.04 | 4.86 | 0.94 | 0.96 | 2.85 | 1.99 |
| 2 | 6.35 | 4.92 | 0.86 | 0.95 | 3.03 | 2.26 |
| 3 | 6.22 | 4.92 | 0.94 | 0.96 | 2.68 | 1.85 |
| 4 | 5.72 | 4.28 | 0.91 | 0.94 | 2.96 | 2.35 |
| **Mean** | **6.14** | **4.67** | **0.92** | **0.95** | **2.91** | **2.14** |

**Table S7**: Results by training seed (A) and AnatCL pretraining fold (B), using the same 140 internal-test participants and 120 ON-Harmony acquisitions. Panel A shows the three primary seeds, with fold 0 for AnatCL. Panel B is a separate checkpoint-sensitivity analysis comparing frozen-Ridge with fine-tuning at seed 0. Mean absolute difference denotes the mean absolute pairwise difference in predicted age across scanners.

| Representation | Full sample 120 acquisitions | Shared scanners 80 acquisitions |
|---|---|---|
| FreeSurfer | 0.95 [0.92, 0.96] | 0.95 [0.93, 0.97] |
| AnatCL–SimCLR | 0.92 [0.89, 0.95] | 0.92 [0.88, 0.95] |
| y-Aware | 0.91 [0.84, 0.94] | 0.93 [0.87, 0.95] |
| AnatCL | 0.92 [0.89, 0.95] | 0.95 [0.93, 0.96] |
| BrainSegFounder | 0.55 [0.53, 0.57] | 0.64 [0.61, 0.66] |
| 3D-Neuro-SimCLR | 0.29 [0.16, 0.39] | 0.34 [0.16, 0.46] |
| NeuroVFM | 0.19 [0.13, 0.24] | 0.20 [0.10, 0.28] |
| Curia-2 | 0.16 [0.11, 0.21] | 0.11 [0.05, 0.16] |
| BrainIAC | 0.11 [-0.02, 0.25] | 0.16 [-0.04, 0.35] |

**Table S8**: Post hoc sensitivity of full-vector dbICC to scanner coverage. Values are dbICC [95% CI]. The full sample contains six acquisitions per participant across the two phase-specific scanner sets (120 acquisitions). The restricted sample retains the same 20 participants on the four shared scanners, NOT1, NOT2, OXF1, and OXF2 (80 acquisitions). Both samples use the first acquisition per participant–scanner pair and the same 1,000 participant-bootstrap resamples.

**Panel A — DKT cortical regions (31 regions × 2 hemispheres × 3 measures = 186 features)**

| | | |
|---|---|---|
| caudalanteriorcingulate | lateralorbitofrontal | posteriorcingulate |
| caudalmiddlefrontal | lingual | precentral |
| cuneus | medialorbitofrontal | precuneus |
| entorhinal | middletemporal | rostralanteriorcingulate |
| fusiform | paracentral | rostralmiddlefrontal |
| inferiorparietal | parahippocampal | superiorfrontal |
| inferiortemporal | parsopercularis | superiorparietal |
| insula | parsorbitalis | superiortemporal |
| isthmuscingulate | parstriangularis | supramarginal |
| lateraloccipital | pericalcarine | transversetemporal |
| | postcentral | |

**Panel B — unique aseg subcortical volumes (15 features)**

| | | |
|---|---|---|
| Brain-Stem | Left-Hippocampus | Right-Hippocampus |
| Left-Accumbens-area | Left-Pallidum | Right-Pallidum |
| Left-Amygdala | Left-Putamen | Right-Putamen |
| Left-Caudate | Left-Thalamus | Right-Thalamus |
| Right-Accumbens-area | Right-Amygdala | Right-Caudate |

**Table S9**: FreeSurfer morphometric feature inventory (201 unique features). **Panel A:** 31 DKT atlas cortical regions, each measured bilaterally for cortical thickness, surface area, and GM volume (186 features). **Panel B:** 15 unique aseg subcortical volumes. The complementary phase-specific Thalamus and Thalamus-Proper columns were coalesced within each hemisphere because they map to the same aseg label.

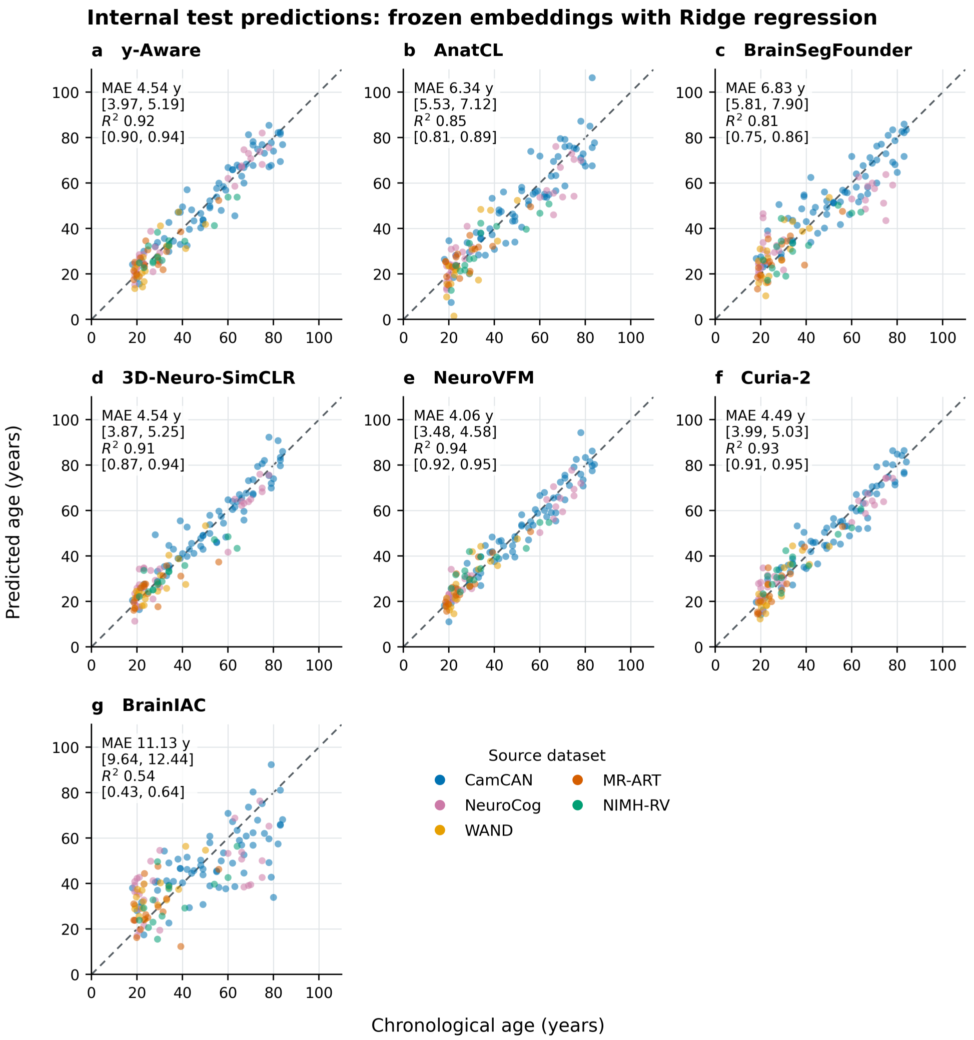


**Fig. S2**: Chronological versus predicted age for the seven frozen-embedding Ridge models in the internal test set. Points are coloured by source dataset, and the dashed line denotes identity. Each panel shows one run, using fold 0 for AnatCL and the sole frozen run for each other model. Each point represents one participant, giving 140 points per model. Insets report MAE and $R^2$ for the displayed run with 95% participant-bootstrap confidence intervals (1,000 resamples).

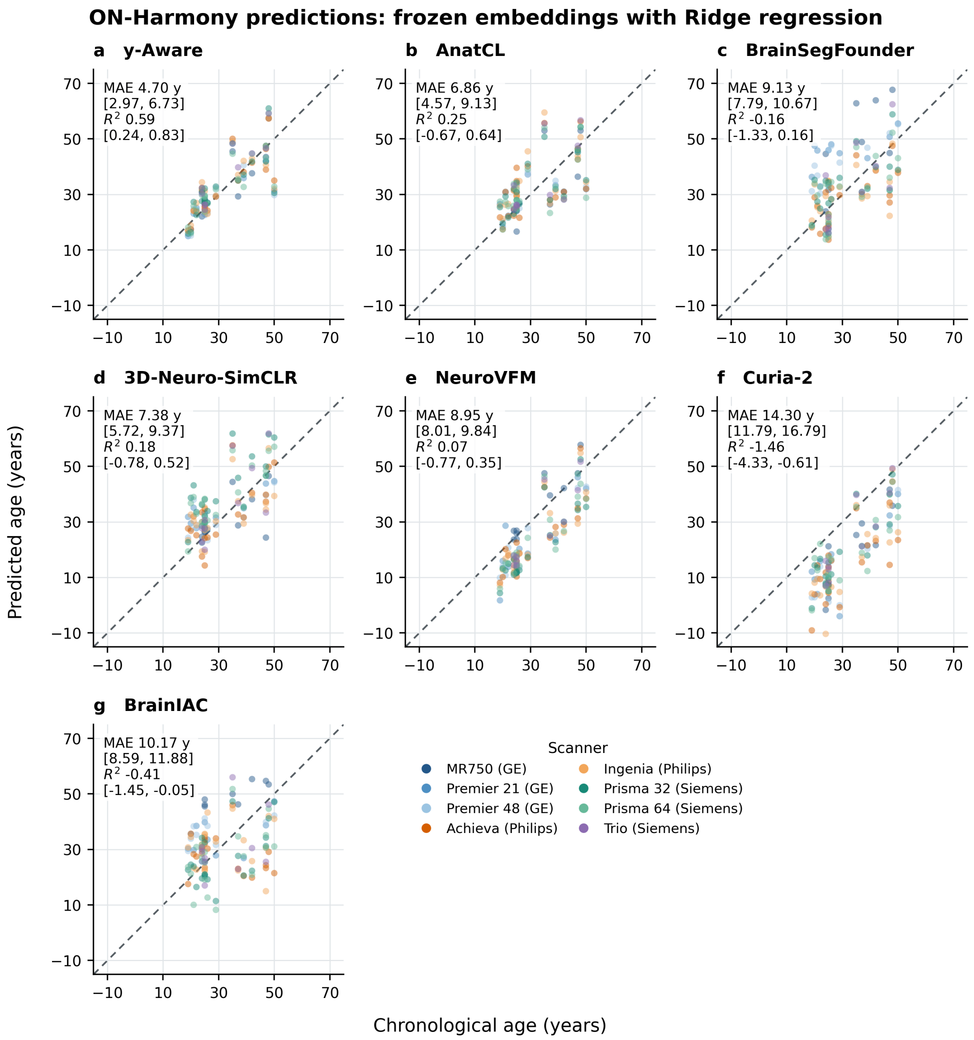


**Fig. S3**: Chronological versus predicted age for the seven frozen-embedding Ridge models in ON-Harmony. Points are coloured by scanner, and the dashed line denotes identity. Chronological age was joined by participant only after label-blind inference. Each panel shows one run, using fold 0 for AnatCL and the sole frozen run for each other model. Each point represents the first acquisition from one participant–scanner cell, giving 120 points per model. Insets report MAE and $R^2$ for the displayed run with 95% participant-bootstrap confidence intervals (1,000 resamples, retaining all six scanner acquisitions per participant).

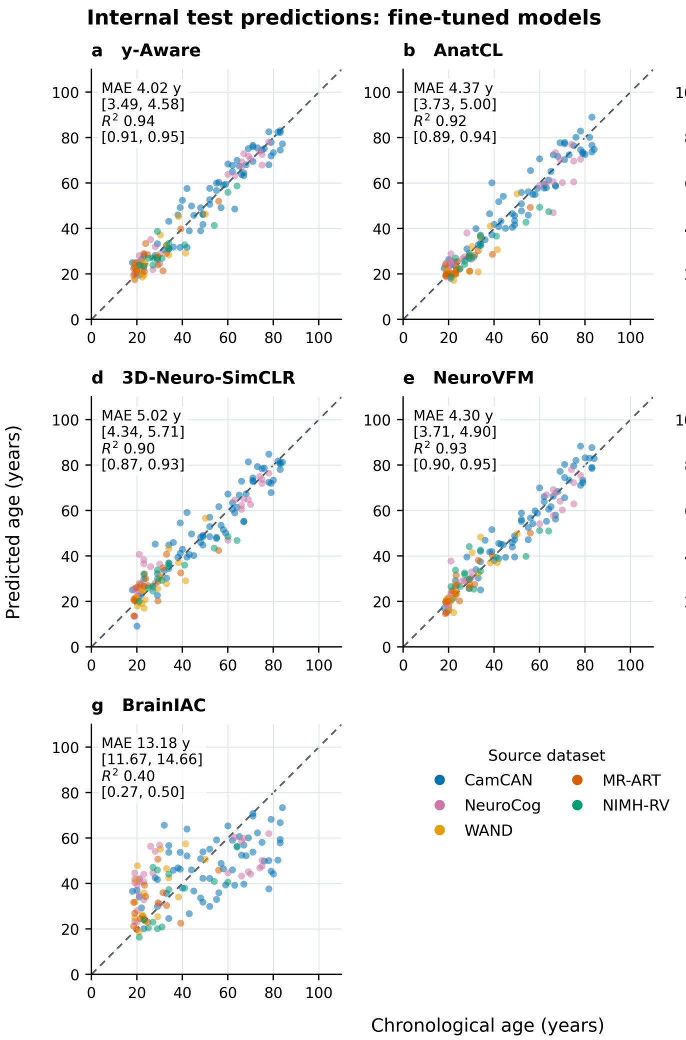


**Fig. S4**: Chronological versus predicted age for the seven fine-tuned models in the internal test set. Points are coloured by source dataset, and the dashed line denotes identity. Each panel shows seed 0, using fold 0 for AnatCL. Each point represents one participant, giving 140 points per model. Insets report MAE and $R^2$ for the displayed run with 95% participant-bootstrap confidence intervals (1,000 resamples). The primary results in the main text average metrics across three seeds for every model.

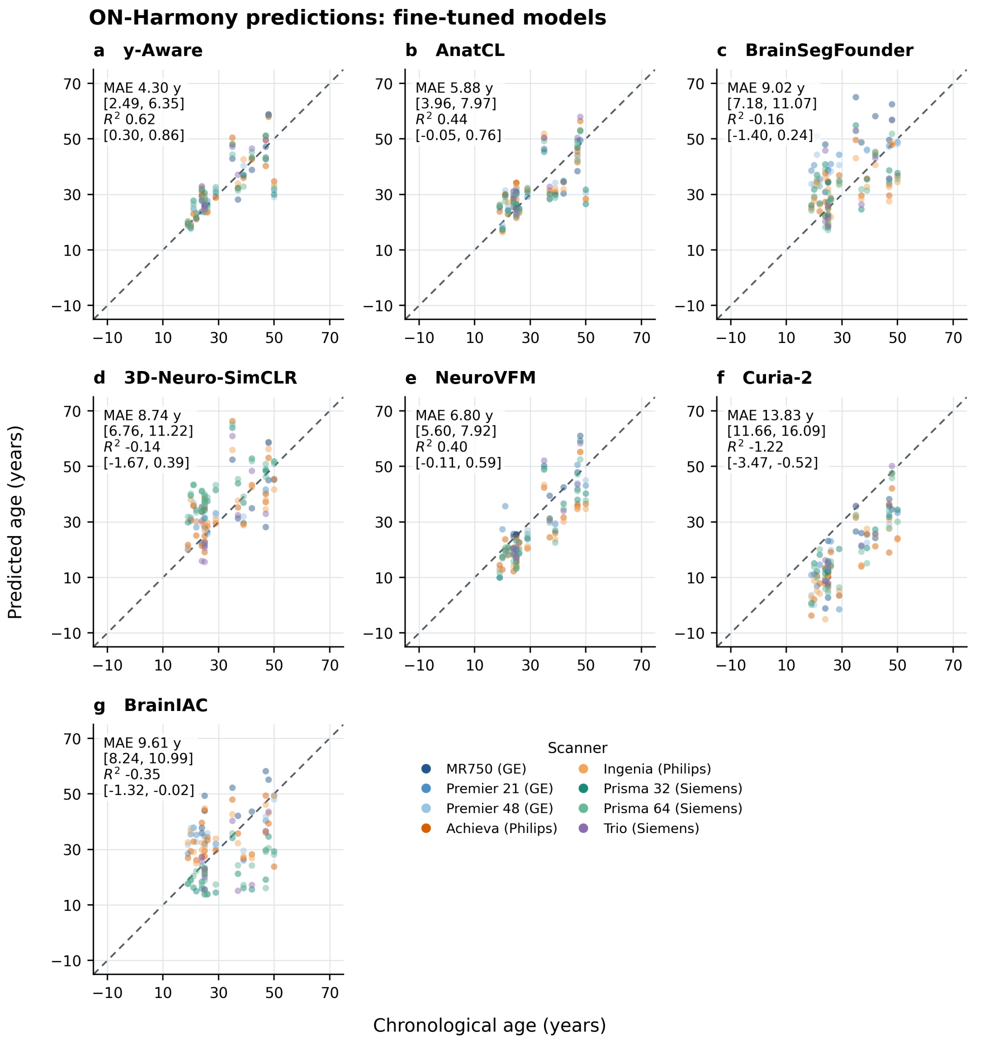


**Fig. S5**: Chronological versus predicted age for the seven fine-tuned models in ON-Harmony. Points are coloured by scanner, and the dashed line denotes identity. Chronological age was joined by participant only after label-blind inference. Each panel shows seed 0, using fold 0 for AnatCL. Each point represents the first acquisition from one participant–scanner cell, giving 120 points per model. Insets report MAE and $R^2$ for the displayed run with 95% participant-bootstrap confidence intervals (1,000 resamples, retaining all six scanner acquisitions per participant). The primary results in the main text average metrics across three seeds for every model.

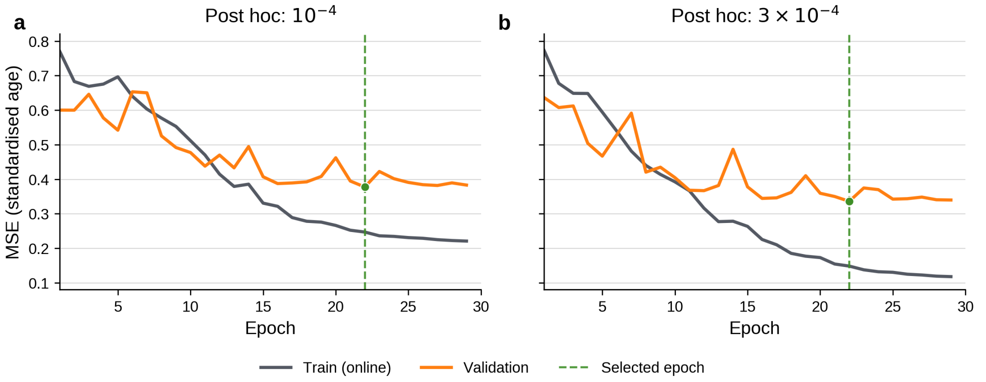


**Fig. S6**: BrainIAC learning-rate sensitivity test using seed 0. The encoder learning rate was increased from $10^{-5}$ in the selected common-grid configuration to $10^{-4}$ (**a**) and $3 \times 10^{-4}$ (**b**), while the age-head learning rate remained $10^{-3}$. Lines show online training MSE and end-of-epoch validation MSE for standardised age. Dashed lines and green points mark the checkpoint selected by validation MAE. Compared with the common-grid validation MAE of 11.88 years, the higher learning rates reduced validation MAE to 10.09 and 9.28 years but produced a wider train–validation separation.
